# The REST (Randomised Evaluation of Sleeping with a Toy or comfort item) Trial: An online, randomised trial of comfort item use on sleep quality in children

**DOI:** 10.64898/2026.02.05.26345698

**Authors:** Simone Lepage, Laura Flight, Nikki Totton, Declan Devane

## Abstract

Sleep is essential for children’s health and development, yet sleep problems are common worldwide. Comfort items such as soft toys or blankets are widely used to promote independent sleep, but their effects have not been evaluated in a randomised controlled trial (RCT). The REST trial emerged from a child-led citizen-science study (The Kid’s Trial) where children co-created and designed the trial. Therefore, this paper had two aims, to assess whether sleeping with a comfort item affected children’s sleep; and to assess the feasibility of conducting an online, child-led citizen-science RCT.

The REST (Randomised Evaluation of Sleeping with a Toy or comfort item) trial was an online two-arm, parallel-group, superiority RCT. Children, aged 7 to 12 years, were randomised (1:1) to either sleep with a self-chosen comfort item (‘Try-it-Out’ group) or refrain from using one (“Wait-and-See” group) for one week. The primary outcome was sleep-related impairment (SRI; PROMIS Pediatric Short Form v1.0 SRI 4a). The secondary outcome was overall sleep quality (Single Item Sleep Quality Scale, SQS). Analyses followed an intention-to-treat principle using mixed-effects models adjusted for baseline measures.

A total of 139 children from 11 countries were randomised (mean age: 9.8 years; 45% female); 101 children (73%) completed post-test measures at one week. The adjusted mean difference (Intervention minus Control) in SRI T-scores was -0.53 (95% CI: -3.40 to 2.34; p = 0.714), equivalent to approximately -0.05 SD on a scale where 10 points = 1 SD. This indicated a trivial effect, well below the minimal important difference (MID) of 3 points. The adjusted mean difference in SQS was 0.28 (95% CI: 0.01 to 0.55; p = 0.040), suggesting a small and uncertain difference in favour of the intervention group. However, this result was not supported in subsequent sensitivity or exploratory subgroup analyses. No adverse events were reported.

Sleeping with a comfort item for one week did not influence sleep-related impairment. A small statistically significant difference in perceived sleep quality was observed in the primary analysis, but was not sustained in the per-protocol analysis. Together, these findings suggest that any benefit of comfort items for sleep is small and uncertain. The trial demonstrated that children can meaningfully engage in online, citizen-science research, supporting the feasibility of child-led RCTs.

**Trial registration:** ISRCTN13756306 (registered 10 January 2025)

## Introduction

Sleep is fundamental to children’s physical, cognitive, and emotional development. Adequate, good-quality sleep supports growth, learning, memory consolidation, and emotion regulation [1–3]. However, sleep problems are common in childhood across many countries and cultures. A 2013 survey of over 900 Chinese children (aged 6 to 14 years) found that 69.3% experienced global sleep disturbances [4]. The 2021 US National Survey of Children’s Health found that between 25% and 50% of over 72,000 children did not get enough sleep, depending on the state in which they lived [5]. In a Finnish study with 1,714 children (aged 4 to 12 years), 21.4% were reported by their parents to have moderate to severe sleep difficulties [6]. Sleep disturbance in children is associated with behavioural difficulties, low mood [6], impaired daytime functioning [2] and increased incidence of obesity [7]. Identifying modifiable factors that may improve sleep is therefore of interest to children, parents, and clinicians alike.

Among the many factors affecting sleep, using sleep aids in the form of comfort items or “transitional” objects (e.g., soft toys, blankets) is a relatively low-cost and easily implementable intervention. The use of comfort items is common in childhood and has been theorised to support emotion regulation during separations and at bedtime [8, 9]. Observational and developmental studies have linked attachment to these objects with self-soothing and coping under stress [10, 11]. Clinical sleep guidance often recommends a comfort item to aid independent sleep in young children [12, 13]. Despite their widespread use, the effects of comfort items on children’s sleep have not been evaluated in a randomised controlled trial (RCT). This gap in the evidence is notable given the potential for a simple, non-invasive and relatively accessible intervention to reduce sleep-related difficulties.

To address this, we developed the REST (Randomised Evaluation of Sleeping with a Toy or comfort item) trial, a two-arm, parallel-group, superiority RCT conducted entirely online. The REST trial was a component of *The Kid’s Trial*, a larger child-led citizen-science study in which children aged 7 to 12 years co-created the trial to learn about RCTs and why they matter. *The Kid’s Trial* guided children through a process of question generation, prioritisation, and planning, where children selected the trial question and planned its design. The question they chose was: *Does sleeping with a comfort item (for example, a soft toy or special blanket) make a difference in how well kids sleep compared with not sleeping with a comfort item?* The details of the citizen-science study that led to the REST trial are reported in our accompanying manuscript, *The Kid’s Trial: Methods and reflections from co-creating and conducting an online, randomised trial with 7 to 12 year old children* [Under Review].

Building on this co-created foundation, the REST trial embodies the principles of citizen science, where participants, in this case, children, contribute directly to the design and conduct of scientific studies. Citizen science offers participatory and experiential learning opportunities that can increase scientific literacy, engagement, and ownership of research [14–16]. While there are excellent examples of this approach extending to RCTs [17, 18], to our knowledge, this is the first fully child-led, online RCT conducted on a global scale, offering a novel model for participatory health research with children.

The REST trial, therefore, had two aims: first, to assess whether sleeping with a comfort item affected children’s sleep; and second, to assess the feasibility of conducting an online, child-led citizen-science RCT. This paper reports the results and implications of the REST trial and discusses the feasibility of conducting child-led citizen-science RCTs.

## Methods

### Ethics and registration

This study received ethical approval on 16 January 2023, from the University of Galway Research Ethics Committee (Ref: 2023.02.014) and was prospectively registered on 10 January 2025, with ISRCTN (ISRCTN 13756306), https://doi.org/10.1186/ISRCTN13756306. This article has been prepared in accordance with the 2025 updated CONSORT guideline for reporting RCTs and the associated explanation [19, 20]. The CONSORT checklist is available in Supporting Information S1 CONSORT Checklist. The accompanying trial protocol can be found as a preprint [21].

### Study design and setting

The REST trial was the RCT component of *The Kid’s Trial*, and was a two-arm, parallel-group, pragmatic, superiority RCT conducted entirely online. Trial materials, including animated explainer videos, guided parents and children through trial processes, consent, randomisation, and outcome assessments. All materials were delivered via a custom-built website (www.TheKidsTrial.ie) with links to QuestionPro survey software [22], which hosted all surveys. Due to the decentralised design, the trial was open to children globally, and all participants joined from home.

The REST trial’s research question originated from the citizen-science study, *The Kid’s Trial*, where children aged 7 to 12 suggested potential research questions. These questions were then narrowed down through two rounds of voting by the participating children. Before the final voting, we searched the Cochrane database, the University of Galway library database, and Google Scholar to confirm that RCTs had not previously addressed the top three candidate questions.

### Participants

Participants were children (7 to 12 years) who met the inclusion criteria as below. Inclusion was confirmed through parental consent and participant assent, obtained via QuestionPro surveys [22]. The REST trial was open to any child worldwide, with no geographical exclusion criteria. However, due to budgetary constraints, we were unable to translate trial materials into other languages; therefore, a good understanding of English was necessary. Below are the inclusion and exclusion criteria.

### Inclusion Criteria

- Children aged 7 to 12 years.
- Proficiency in English sufficient to understand trial materials.
- Access to the trial’s online platform.
- Parental consent for participation.

### Exclusion Criteria

- Inability to understand and provide assent.

### Participant recruitment and demographics

Recruitment was mainly conducted online via social media (X, Facebook, Instagram, TikTok, Mastodon, and LinkedIn) and email campaigns. The study was also listed on two websites: the European Citizen Science Platform [23] and Children Helping Science [24], which connect the public with research opportunities. Recruitment efforts were further expanded by testing paid Google ads for two weeks and running a three-month paid Facebook campaign. Offline recruitment methods included national radio interviews and distributing flyers to schools, markets, play centres, and by collaborating with children’s advocacy groups in countries where we had no existing networks.

Recruitment was passive, with children and parents initiating contact through email, social media, or the trial website. The website displayed the Participant Information Leaflets (PILs), called the ‘Parents’ Information Flipbook’ and the ‘Children’s Information Flipbook’. Participants were informed through these flipbooks and during the consent and assent processes that they could withdraw at any time without consequences, and their data could be removed before data analysis began. Enrolment was rolling, so each participant’s trial start and end dates depended on their date of joining.

Due to the participants’ age, parents were asked to provide their child’s demographic details. To meet the inclusion criteria, the child’s age was required, while gender, country of residence, and ethnicity were optional to assess the trial’s inclusivity and geographical coverage.

### Randomisation and blinding

To join the REST trial, children clicked a ‘Join the trial here!’ button on the website, which directed them to a QuestionPro [22] survey where, after entering demographic and baseline sleep data, they were randomised to either the intervention (‘Try-it-Out’) or control (‘Wait-and-See’) group. Randomisation was computer-generated within the QuestionPro survey software [22]. Due to the constraints of the software, group allocation was performed using simple randomisation (1:1). Participants could not predict their group allocation; however, both they and the researchers became aware of the allocation after randomisation. Neither researchers nor participants had any influence over the allocation. Due to resource constraints, it was not possible to blind researchers to participants’ group allocation, and because of the nature of the intervention, it was impossible to blind participants to their group allocation. Participants were instructed to begin their trial the day after they were randomised to their treatment group.

### Interventions

The parameters for both the intervention and control arms were decided prior to the start of the trial through a vote by children participating in *The Kid’s Trial* [Under Review].

Participants allocated to the intervention arm, the ‘Try-it-Out’ group, were asked to select a comfort item, defined as any item they believed might bring them comfort (e.g., a soft toy or special blanket), and to sleep with it for seven consecutive nights. They were instructed to use the same item throughout the trial, sleep in their usual bed, and start using the item at the beginning of their normal bedtime routine, while continuing their usual routines otherwise. Bedtime routines were described as any activities children typically perform each night to prepare for sleep. For example, if they read every night before bed, they were asked to begin using their comfort item at that time. Children’s ‘usual beds’ were defined as any bed in which they regularly slept.

Participants in the control arm, the ‘Wait-and-See’ group, were asked to avoid sleeping with any comfort item for seven consecutive nights. They were also instructed to sleep in their usual bed and maintain their regular bedtime routine.

### Outcomes

To test whether sleeping with a comfort item affected children’s sleep, we used two self-reported measures. In the ‘Planning the trial!’ phase of *The Kid’s Trial*, children voted on how sleep should be assessed. They chose sleep-related impairment (SRI) as the primary outcome and overall sleep quality as the secondary outcome.

Participants completed outcome measurement questions at baseline when they joined the trial, and again eight days after randomisation, at post-test, when their trial was finished. At post-test, participants’ parents received an email with a link to the outcome measurement survey specific to their allocated group. The post-test outcome survey asked the same questions that participants were asked at baseline, with the addition of group-specific intervention adherence questions (available in Supplementary Information S2 Outcome Questionnaires). To enhance retention and reduce missing data, parents received two email reminders if their children had not completed their questionnaire at post-test. Email reminders were sent on day 10 and day 13 post-randomisation.

### Primary outcome

To measure SRI, we used the Patient Reported Outcomes Measurement Information System (PROMIS) Pediatric Short Form v1.0 SRI 4a [25]. The instrument consists of four short questions concerning daytime sleep-related impairment, with participants selecting one of five options: ‘Never’, ‘Almost Never’, ‘Sometimes’, ‘Almost Always’, or ‘Always’. Response options are scored from 1 to 5, except for the final item, which is scored from 1 to 4 because ‘Almost Always’ and ‘Always’ are both scored as 4 points [26]. The total raw score, therefore, ranges from 4 to 19, with higher scores indicating greater impairment. Each question’s raw score contributes to the total, which is then converted to a standardised T-score using the PROMIS scoring manual conversion table [26, 27]. Further PROMIS SRI scoring details are available in the statistical analysis plan (SAP). The SAP was made available at the time of the protocol preparation and can be found in an Open Science Framework (OSF) repository [28], and is included in Supporting Information S3 SAP.

### Secondary outcome

To measure overall sleep quality, we used the Single Item Sleep Quality Scale (SQS) instrument [29]. The SQS consists of one question, asking participants to rate their overall sleep quality over the previous seven days on a categorical scale from 0 to 10, where 0 is ‘terrible’, 1-3 is ‘poor’, 4-6 is ‘fair’, 7-9 is ‘good’, and 10 is ‘excellent’. As per the instrument guidance, the 0-10 ratings are collapsed into five categories, coded to a 1-5 score, with higher scores indicating better sleep quality [29]. No further conversion was needed to score this measure. Primary and secondary outcome questionnaires are available in Supporting Information S2 Outcome Questionnaires.

### Covariates

We used baseline measurements of the outcome being analysed, baseline comfort item use, age, and gender as fixed covariates. The country of residence served as the random effect, as detailed in the SAP (S3 SAP). However, because several countries had only one participant, small countries were collapsed into regional categories to ensure model convergence and stable variance estimation: Austria, Estonia, and Germany were grouped as “Europe”; India and Pakistan were grouped as “South Asia”; Australia and Mexico were retained as single-country groups. This collapsed country variable was carried throughout all analyses.

### Adherence and treatment integrity

Post-test, participants were asked whether they followed their group’s instructions, using a Likert scale ranging from ‘Never’ to ‘Always’, with options of ‘1-2 nights’, ‘3-4 nights’, and ‘5-6 nights’. Adherence depended on group allocation. If a participant adhered to their group’s treatment instructions for fewer than 5-6 nights, they were considered non-adherent and asked why they deviated from their group’s instructions.

### Sample size

The target sample size was 292 (146 per arm), providing 80% power (two-sided α=0.05) to detect a minimally important difference (MID) of 3 T-score points on SRI [30], assuming a standard deviation (SD) of 10, correlation between repeated measures r=0.5, and 10% attrition, as specified in the SAP (S3 SAP).

### Statistical analyses

We used R (version 4.3.1) [31] for all analyses. All R coding scripts are available in Supporting Information S4 R Markdown Scripts. The primary outcome, SRI, was analysed using participants’ T-scores (lower scores indicating less sleep-related impairment), and the secondary outcome, overall sleep quality, was analysed using SQS raw scores (higher scores indicating better sleep quality).

### Primary analysis (ITT)

Our primary analyses followed an Intention-to-Treat (ITT) principle. For each outcome, we analysed the post-test score using a mixed-effects model with the treatment group serving as the main predictor. Our fixed covariates were the baseline value of the outcome being analysed, baseline comfort-item use (Never/Sometimes/Always), age, and gender. We used the country (collapsed country, where applicable) of residence as a random effect. We report adjusted means by group, along with the adjusted mean difference (Intervention minus Control), 95% confidence intervals (CIs) and two-sided p-values.

#### Model checks

Model assumptions were checked using residual-versus-fitted, normal Q–Q plots, and histograms of the residuals. The estimated country random-effect variance was assessed to ensure its inclusion was appropriate.

### Exploratory subgroup analyses

We assessed whether the treatment effect varied by baseline comfort-item use, age group (7–9 vs. 10–12), and gender by adding group-by-subgroup interaction terms to the primary model and reporting a global interaction p-value for each. Subgroups are presented with the adjusted mean difference and confidence intervals with less reliance on the p-value due to the exploratory nature. As they are exploratory, they were not adjusted for multiple comparisons.

### Sensitivity analyses

#### Per-protocol analyses

We repeated the primary mixed-effects model in a per-protocol (PP) subset, restricted to participants who adhered to their allocated treatment as intended (≥5 nights consistent with the assigned treatment).

#### Multiple imputation analyses

Because ∼27% of post-test outcomes were missing, we performed multiple imputation (MI) under a Missing-At-Random (MAR) assumption. The imputation model included the post-test outcome, baseline outcome, treatment group, baseline comfort-item use, age, gender, and country of residence. Each imputed dataset was analysed with the primary model, and estimates were combined using Rubin’s rules [32] to report adjusted mean differences with 95% CIs. For transparency, we present both observed and multiple imputation estimates side by side.

### Adverse events and stopping guidelines

The trial question, “Does sleeping with a comfort item, for example, a soft toy or special blanket, make a difference to how well kids sleep compared with not sleeping with a comfort item?” was a low-risk intervention that reduced the likelihood of any harm associated with the trial. Given the short duration of the trial, there were no stopping guidelines. Additionally, we informed participants they should stop their trial if they felt upset.

## Results

The REST trial opened on 13 January 2025 and closed on 15 September 2025. We aimed to recruit 292 children, but did not achieve this goal. A total of 139 children participated in the trial and were randomly assigned to one of two treatment groups (n = 72 control, n = 67 intervention). Of these, 101 participants submitted outcome data post-test. Attrition was similar between groups: Intervention 17/67 (25%) vs Control 21/72 (29%). The CONSORT participant flow diagram is displayed in Fig 1.

**Fig 1.**
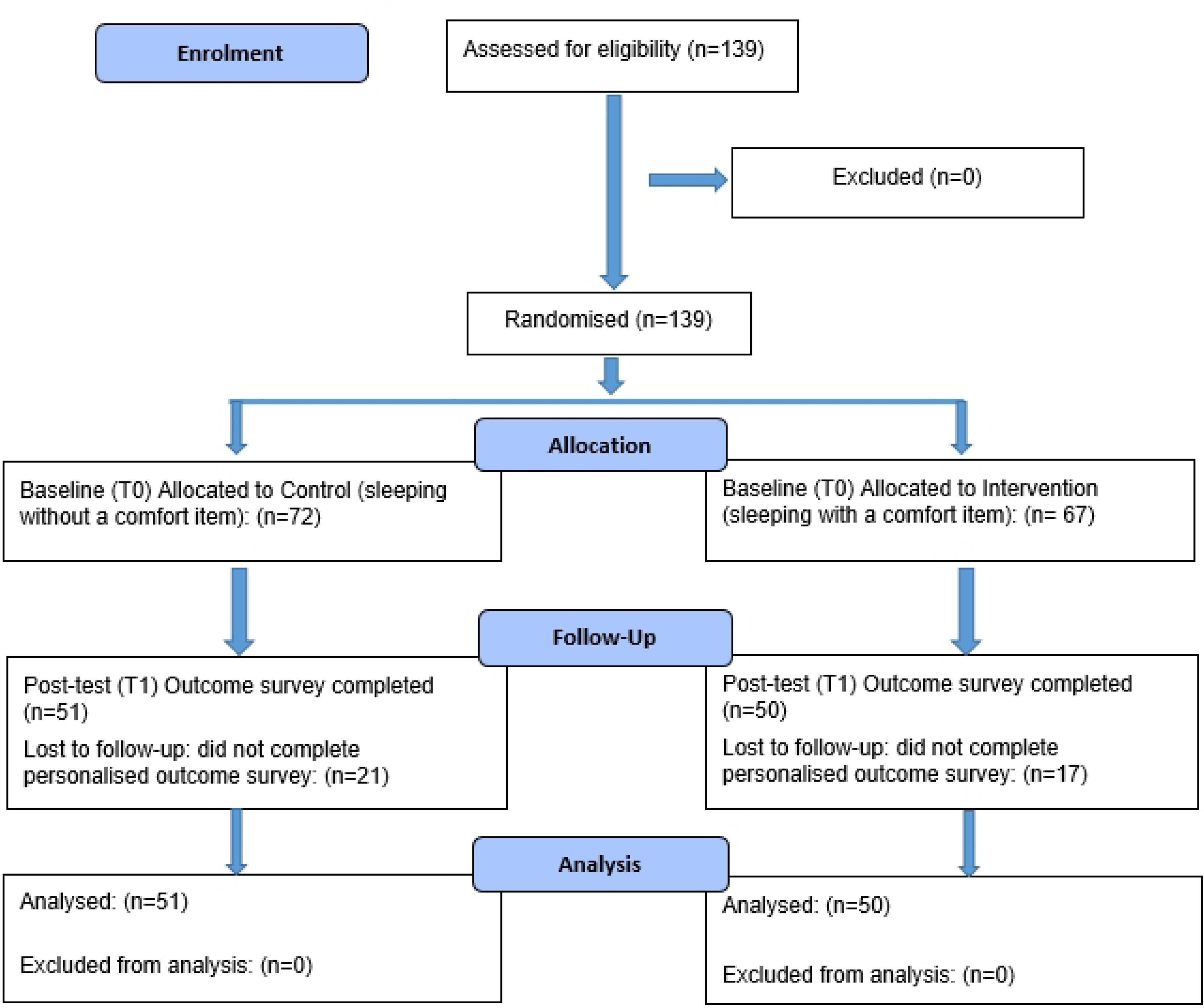
CONSORT 2025 Flow Diagram for the REST trial.

### Baseline characteristics

The mean age of participating children was 9.76 years (SD 1.60) and was similar across arms. The participants comprised 55% males and 45% females. Children from 11 different countries participated in the REST trial, with the majority residing in Kenya (52%) or Ireland (34%). Of the 135 participants who reported ethnicity, 45% chose ‘Black/Black Irish’ and 38% chose ‘White’. The most common category at baseline for comfort-item use was ‘Always’ (45%), which was higher in the intervention arm than in the control (52% vs. 38%). Baseline outcome measurements were similar between arms. The overall mean PROMIS SRI T-score was 51.38 (SD 7.94), which was similar across both treatment groups: Control (51.22) and Intervention (51.55). The overall SQS mean was 3.76 (SD 0.81), which was also similar across treatment groups (Control: 3.79; Intervention: 3.72). All baseline characteristics are shown in Table 1.

**Table 1.**
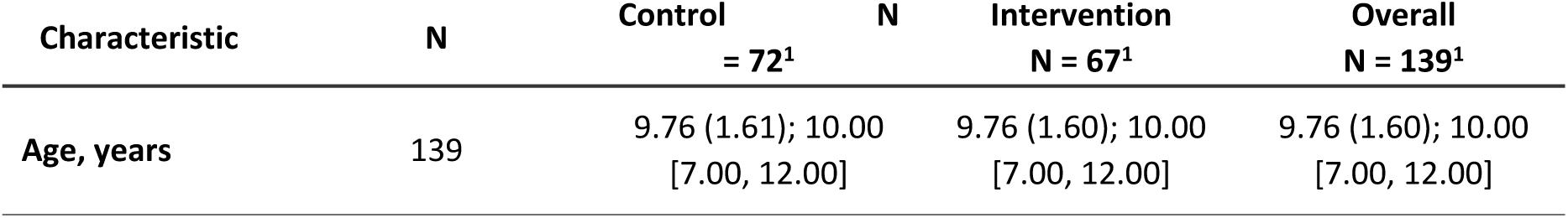
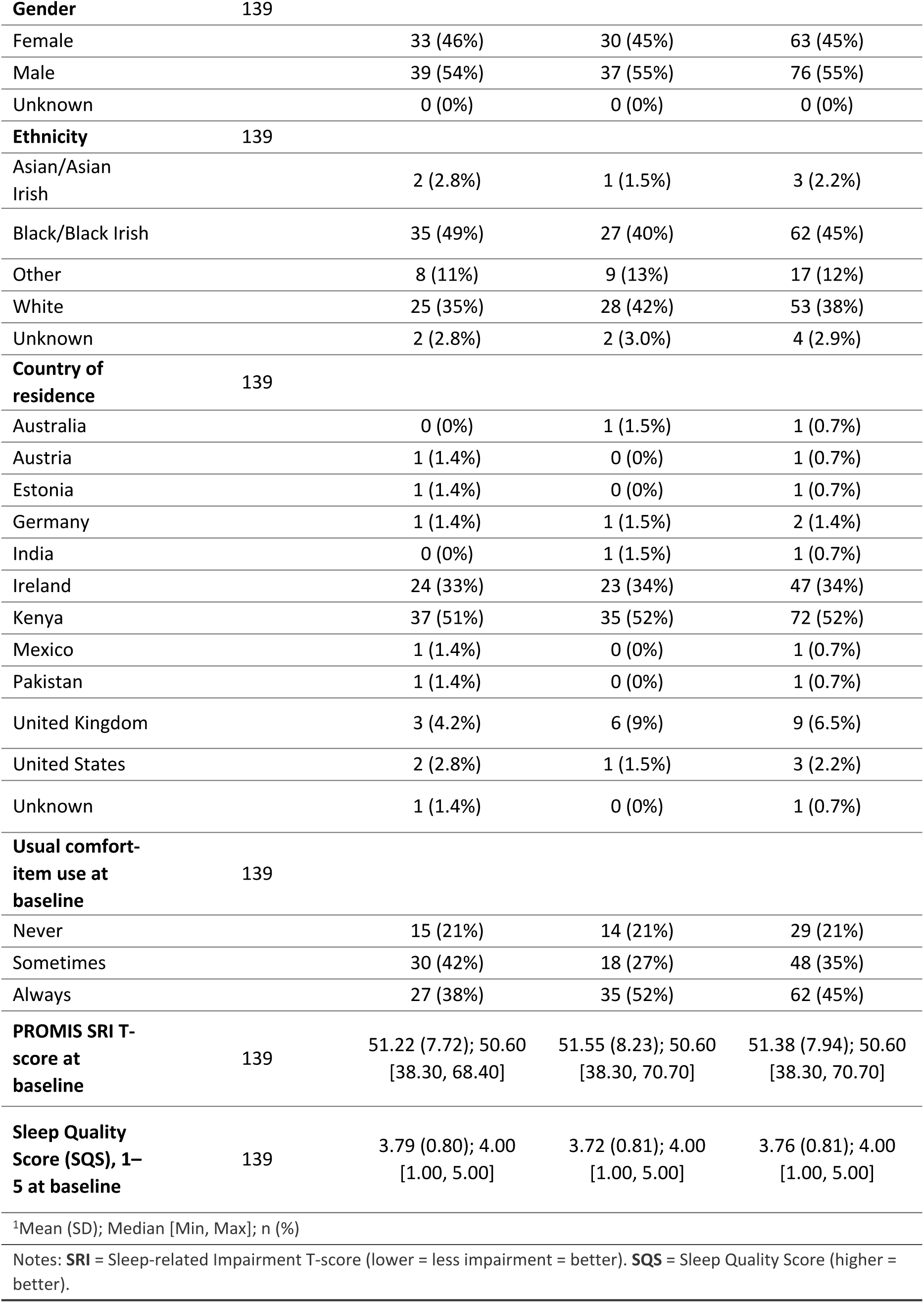
Baseline characteristics. Values are presented as mean (SD) and median [IQR] for continuous variables, and as n (%) for categorical variables.

### Primary analysis results

We analysed all randomised participants with post-test outcomes (Control, n = 51; Intervention, n = 50). The post-test adjusted mean SRI T-score was 47.20 (95% CI: 45.10 to 49.30) in the intervention group, and 47.73 (95% CI: 45.77 to 49.68) in the control group (Fig 2). The adjusted mean difference was -0.53 (95% CI: -3.40 to 2.34; p = 0.714), indicating no statistically significant difference in SRI post-test between groups.

**Fig 2.**
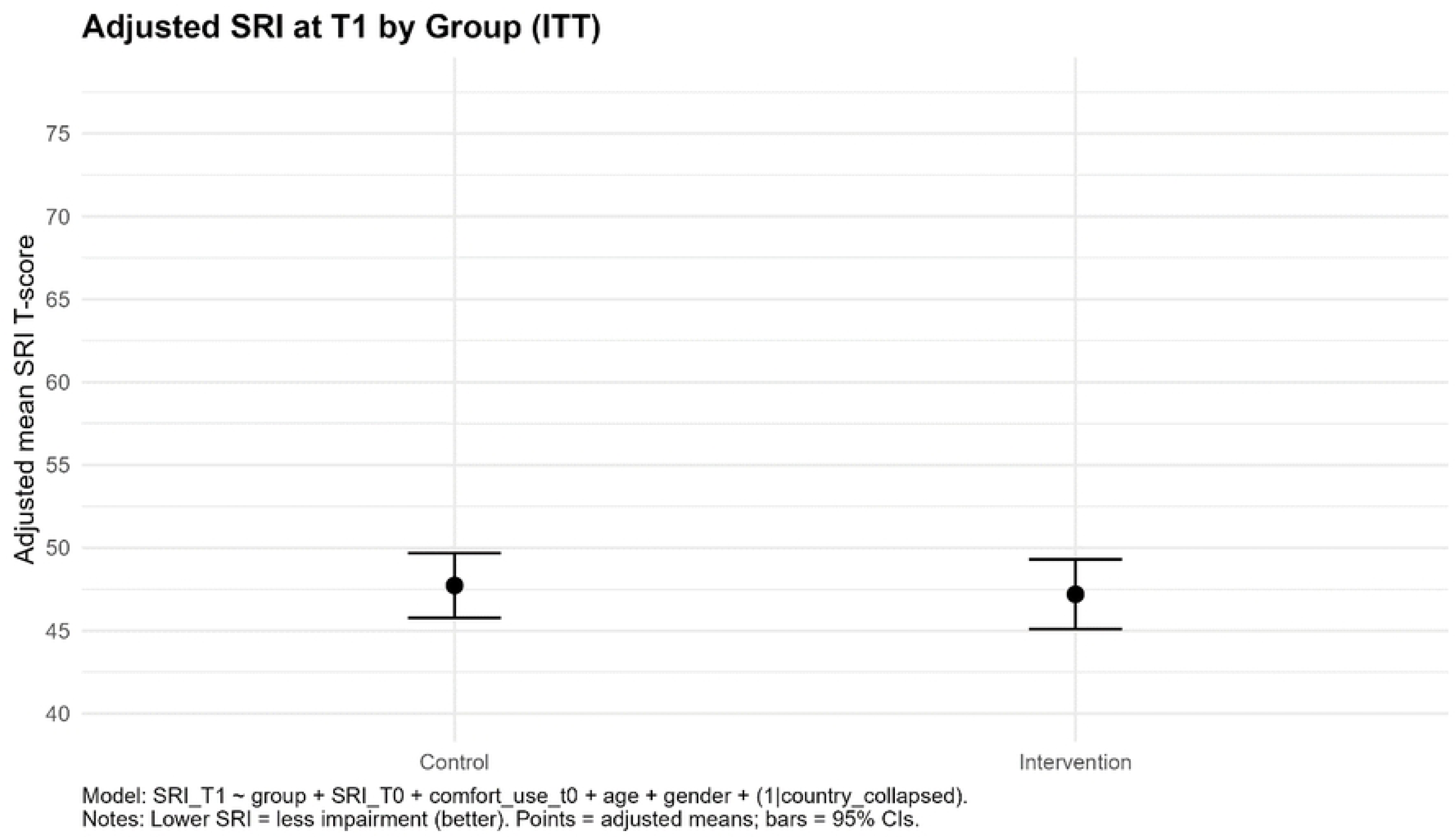
Adjusted SRI at post-test by treatment group (ITT) (means with 95% CIs).

The post-test adjusted mean SQS score was 3.99 (95% CI: 3.79 to 4.19) in the intervention group, and 3.71 (95% CI: 3.52 to 3.89) in the control group (Fig 3). The adjusted mean difference between the groups was 0.28 (95% CI: 0.01 to 0.55; p = 0.0405), representing a statistically significant difference between groups, indicating that those who slept with a comfort item (intervention group) had better overall sleep quality. Table 2 presents the adjusted mean differences in both outcomes (Intervention minus Control) along with their corresponding p-values.

**Fig 3.**
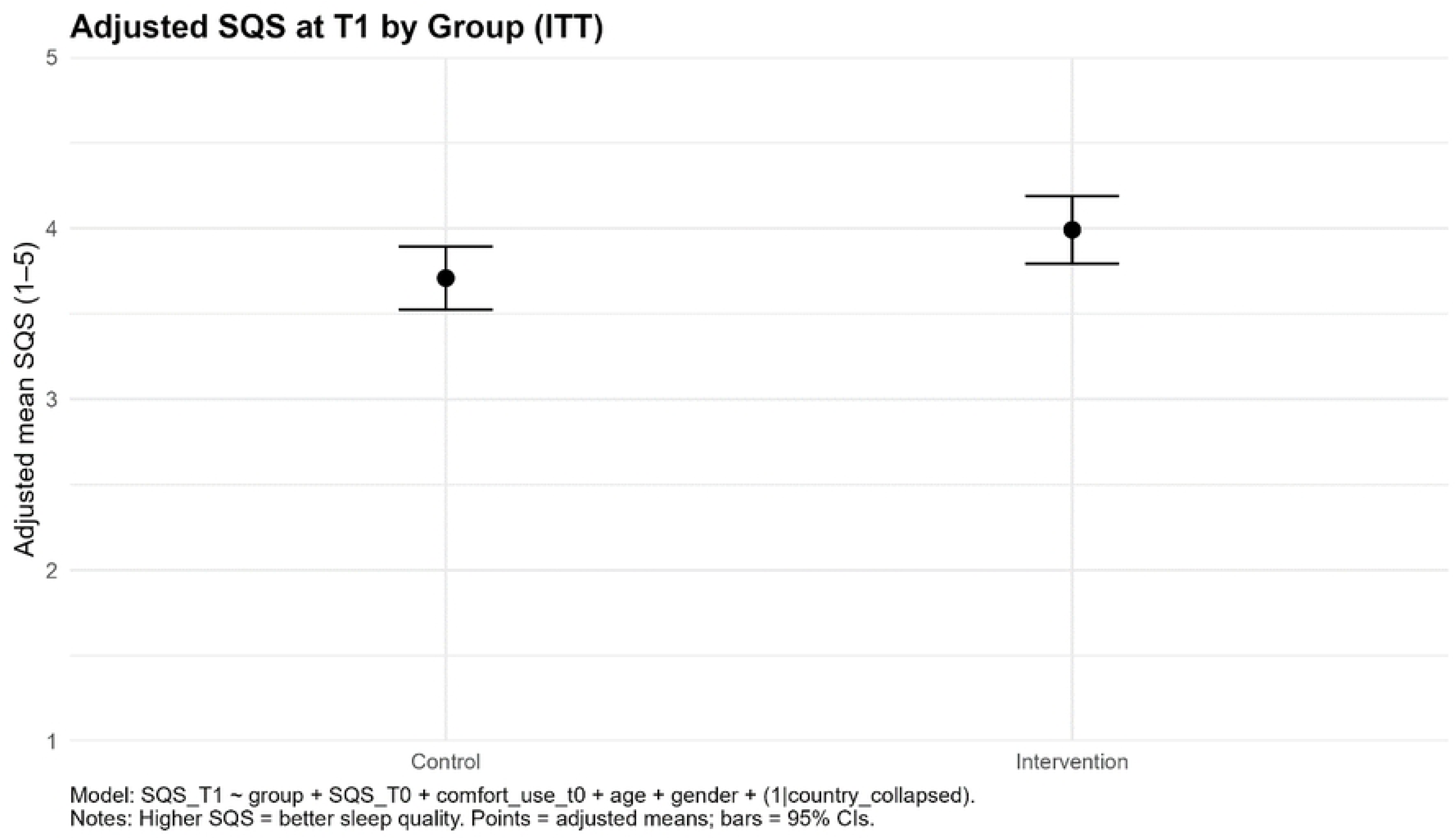
Adjusted overall sleep quality at post-test by treatment group (ITT) (means with 95% CIs).

**Table 2.**
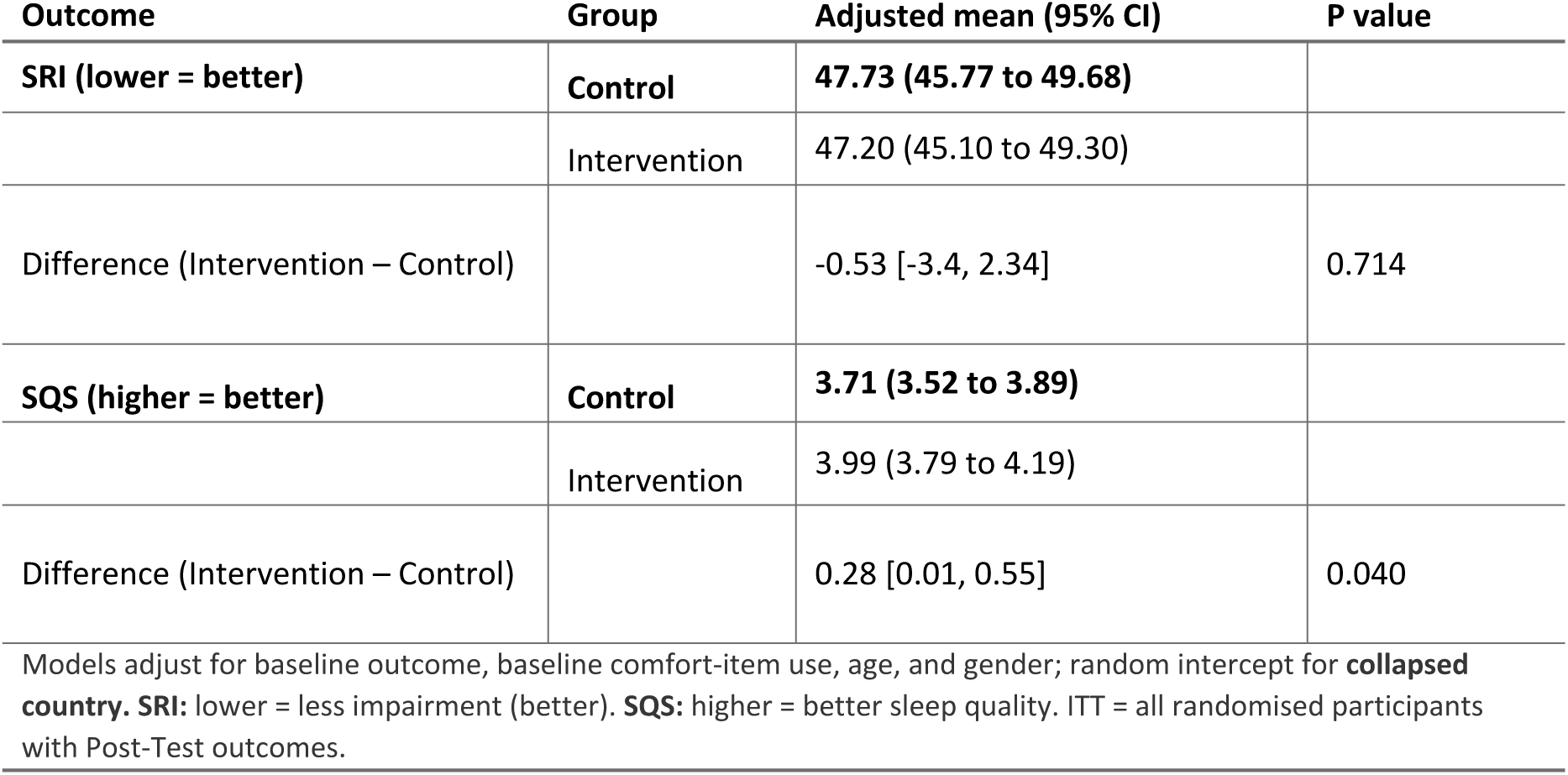
Primary analysis (ITT): Adjusted means and adjusted differences at post-test.

All diagnostic plots showed that the models fit the data adequately. The country random-intercept variance was near zero (‘singular’ fit), implying little between-country clustering, and no refitting was required.

### Exploratory subgroup analyses

Table 3 summarises the exploratory subgroup analyses for both outcomes. For SRI, adjusted treatment effects were small and did not differ meaningfully by baseline comfort-item use, age group, or gender. For SQS, the adjusted mean difference was largest among children who ‘Always’ used a comfort item at baseline (0.65 [0.25, 1.06]), with smaller differences or null differences among those who ‘Sometimes’ (0.18 [-0.29, 0.65]) or ‘Never’ used one (-0.24 [-0.77, 0.29]). Apparent differences by age or gender were small with wide, overlapping CIs, providing no consistent evidence of effect modification.

**Table 3.** Summary of subgroup analyses for adjusted mean difference at post-test.

| Outcome | Moderator | Level | N<br>(Intervention) | N<br>(Control) | Adjusted mean<br>difference<br>(95% CI) | p-value | Global<br>interaction p |
| --- | --- | --- | --- | --- | --- | --- | --- |
| SQS | Age group<br>(years) | 10–12 | 31 | 31 | 0.19 [-0.15,<br>0.53] | 0.268 |  |
| SQS | Age group<br>(years) | 7–9 | 19 | 20 | 0.42 [-0.00,<br>0.85] | 0.053 | 0.397 |
| SQS | Baseline<br>comfort-item use | Always | 29 | 16 | 0.65 [0.25,<br>1.06] | **0.002** |  |
| SQS | Baseline<br>comfort-item use | Never | 10 | 14 | -0.24 [-0.77,<br>0.29] | 0.379 | **0.031** |
| SQS | Baseline<br>comfort-item use | Sometimes | 11 | 21 | 0.18 [-0.29,<br>0.65] | 0.450 |  |
| SQS | Gender | Female | 24 | 24 | 0.21 [-0.17,<br>0.59] | 0.276 | 0.592 |
| SQS | Gender | Male | 26 | 27 | 0.35 [-0.02,<br>0.73] | 0.066 |  |
| SRI T-score | Age group (years) | 10–12 | 31 | 31 | -0.65 [-4.40, 3.11] | 0.734 |  |
| SRI T-score | Age group (years) | 7–9 | 19 | 20 | -0.26 [-4.87, 4.35] | 0.911 | 0.899 |
| SRI T-score | Baseline comfort-item use | Always | 29 | 16 | -2.64 [-6.93, 1.64] | 0.224 |  |
| SRI T-score | Baseline comfort-item use | Never | 10 | 14 | 4.48 [-1.25, 10.22] | 0.124 | 0.136 |
| SRI T-score | Baseline comfort-item use | Sometimes | 11 | 21 | -1.50 [-6.60, 3.60] | 0.560 |  |
| SRI T-score | Gender | Female | 24 | 24 | -0.43 [-4.37, 3.77] | 0.885 | 0.872 |
| SRI T-score | Gender | Male | 26 | 27 | -0.76 [-4.76, 3.25] | 0.708 |  |

Difference is **Intervention - Control**. Models adjust for baseline outcome, baseline comfort-item use, age, and gender, with a random intercept for **collapsed country**.
For SRI, lower scores = less impairment (better). For SQS, higher scores = better sleep quality.
Global interaction p is shown once per (Outcome x Moderator).

### Sensitivity analyses

We repeated the primary mixed-effects model in the per-protocol group. Among children who followed their assigned instructions for at least five nights, the post-test adjusted mean difference for SRI was -0.96 points (95% CI, -4.37 to 2.44; p = 0.574). The CI is wide and crosses zero, indicating uncertainty in the estimate. This result is larger than the ITT estimate (-0.53) but remains well below the MID of 3 points, suggesting that the mean effect is not clinically meaningful [30]. The similarity between the ITT and per-protocol results indicates that non-adherence did not obscure a substantial treatment effect.

For SQS, the per-protocol point estimate was 0.18 points (95% CI: -0.165 to 0.53; p = 0.296). This is similar in size to the ITT estimate (0.28) but is no longer statistically significant. This suggests that any benefit in the perceived sleep quality is small, and the loss of statistical significance is likely due to the smaller sample size in the per-protocol analysis.

Approximately 27% of randomised participants did not provide post-test outcomes; therefore, we performed multiple imputation, including treatment group, baseline outcome, baseline comfort-item use, age, gender, and country. The pooled SRI treatment effect from multiple imputation was close to the complete-case estimate, with a 95% CI crossing zero, indicating no material change in inference (Fig 4). Taken together, multiple imputation did not change the conclusion. For SQS, MI produced results consistent with the complete-case analysis (Supporting Information S5 Observed vs. multiple imputation SQS at post-test).

**Fig 4.**
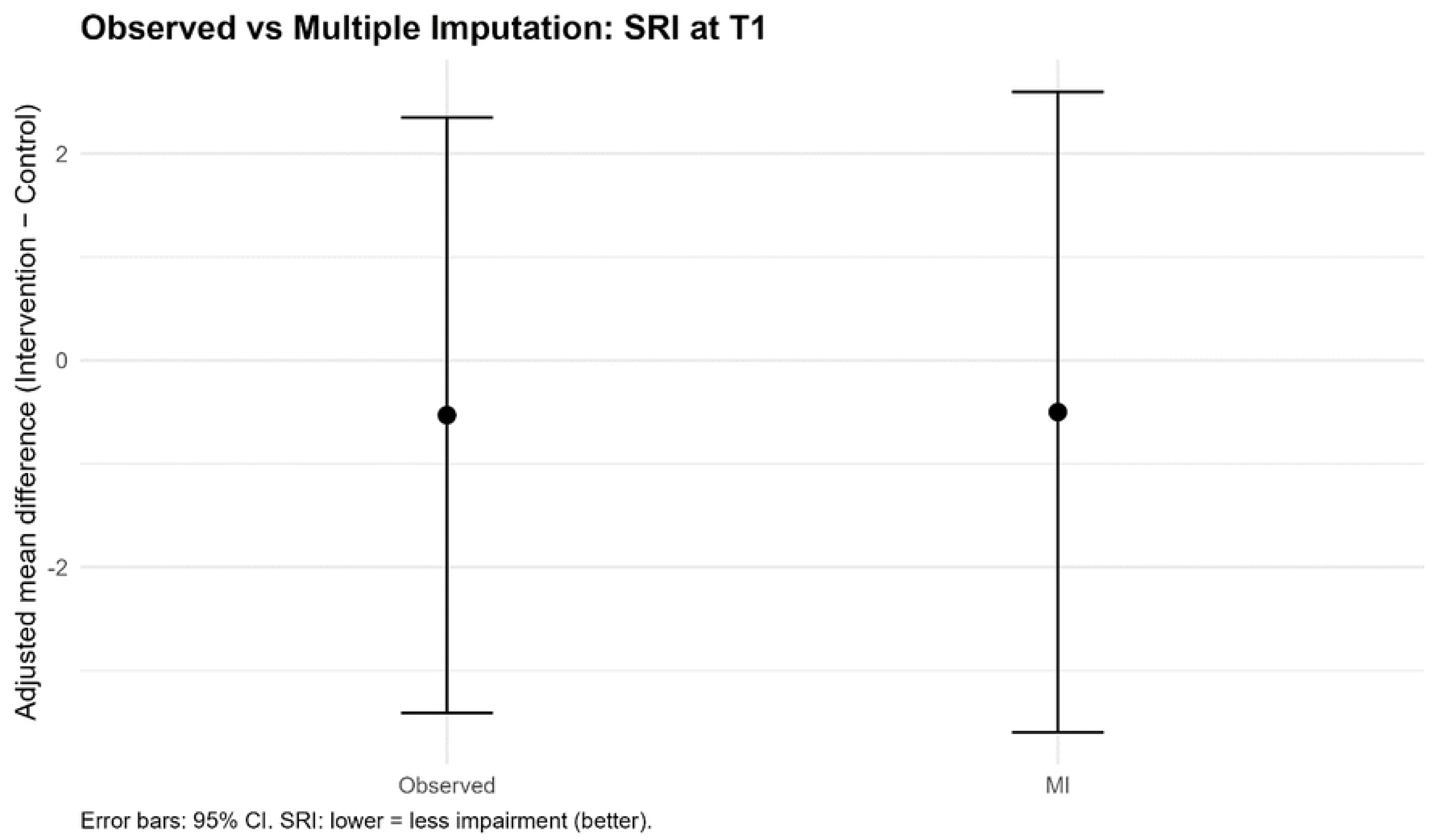
Observed versus multiple imputation adjusted mean difference in SRI.

### Adverse Events

Neither parents nor children reported any adverse events during or after the trial.

## Discussion

In this online, child-led RCT, sleeping with a comfort item for one week did not improve sleep-related impairment on the PROMIS SRI primary outcome. The adjusted mean difference was −0.53 T-score points, which is well below the MID of 3 points. Because SRI is a T-metric with SD 10, the SRI effect corresponds to an approximately 0.05 SD change, which is trivial. For the secondary SQS outcome, the adjusted mean difference was 0.28, a small effect that was not consistent across the per-protocol analysis. These results indicate no clinically important benefit to SRI and a small, uncertain difference in SQS.

Subgroup analyses were exploratory, unadjusted for multiplicity, and should be interpreted cautiously. They provided limited evidence that treatment effects varied by participant characteristics. A modest pattern was observed for SQS among children who consistently used a comfort item, but this requires confirmation in future research. However, any patterns are hypothesis-generating and would require confirmation in future, adequately powered studies. We performed exploratory per-protocol analyses to assess whether the ITT findings remained robust when examined among children considered adherent to their treatment. In these analyses, the results were similar in direction to those in the ITT: SRI again showed no statistically significant difference, and the SQS estimate remained positive but was no longer statistically significant. Taken together, the pattern across ITT and per-protocol analyses reduces confidence that the SQS difference reflects a reproducible effect.

Approximately 27% (38/139) of the randomised participants did not submit their post-test outcome data, which exceeded the 10% SAP threshold. Multiple imputation was used to reduce potential bias from this missing data. The multiple imputation analyses yielded pooled estimates similar to the complete-case results for both outcomes, indicating that the conclusions of the primary analyses remained unchanged after accounting for missing data.

Across the ITT, per-protocol, and multiple imputation analyses, we did not detect a clinically important improvement in SRI. The modest SQS gains seen in the ITT analysis were not supported by the per-protocol analyses, indicating that any benefit for sleep quality remains uncertain and likely modest.

Inconsistent bedtime routines are associated with an increased risk of children’s sleep disturbances [33]. Both children and parents have previously noted that bedtime routines, including comfort items, can help reduce barriers to sleep and promote self-soothing [34]. While comfort items may be part of a bedtime routine, our results suggest that they do not provide a benefit to sleep-related impairment over one week.

Although the trial was not powered to detect subgroup differences, exploratory subgroup analyses suggested that the effect of sleeping with a comfort item on sleep quality might differ by baseline comfort-item use, with the largest apparent benefit among children who already used a comfort item at baseline. These findings should be interpreted with caution. However, they may help guide future research. Future trials could examine whether comfort items are most effective for children who already have a strong attachment to these objects using adequately powered designs and pre-specified subgroup hypotheses. Given the high proportion who reported ‘Always’ using a comfort item at baseline, ceiling effects are possible.

### Feasibility and engagement

Beyond the clinical question this RCT aimed to answer, the REST trial demonstrated that primary-school-aged children can meaningfully take part in a citizen-science trial, potentially improving their health literacy and understanding of evidence creation. The very choice of engaging with the REST trial illustrates children’s and parents’ interest in their own health and their ability to participate in research rigorously. Participation likely increased awareness of sleep hygiene and health literacy, a critical public health goal that ideally begins in childhood [35]. These feasibility observations are encouraging but are separate from the null primary effect and should be interpreted as such.

### Strengths and limitations

The REST trial successfully implemented a child-led RCT with a transparent, pre-registered protocol and a priori SAP. The fully decentralised approach allowed for global participation and reached children who would not have been able to participate if we had limited it to a local population.

The REST trial also had limitations. The study was underpowered. Recruitment was challenging despite the multiple strategies employed. The listing of the trial on the Children Helping Science [24] website only went live in the last two weeks of the trial, likely limiting enrolment via that avenue. Outcomes were self-reported, introducing possible response bias. The intervention lasted only a week to minimise the burden on participants and families, limiting observation of longer-term effects. Blinding was not possible due to the nature of the intervention and budgetary constraints.

Most participants were from Kenya and Ireland, and a high proportion reported ‘Always’ using a comfort item at baseline. As a result, effect estimates may not generalise to settings where comfort item use is less common or carries different meanings. Non-adherence in the control group was also possible, as it cannot be confirmed whether children in that arm refrained from using comfort items. Finally, the online nature of the trial inherently excluded children who lacked internet access.

For families and clinicians, comfort items appear unlikely to reduce sleep-related impairment over one week in children aged 7 to 12 years. If families choose to use them, expectations should be modest. Future trials should extend follow-up, incorporate ordinal models for SQS, pre-specify fewer subgroups, consider actigraphy or sleep diaries as objective or corroborating measures, and explore a complier-average causal effect analysis to account for adherence.

## Conclusion

In this child-led, online RCT, sleeping with a comfort item for one week did not reduce sleep-related impairment. A small statistically significant difference in perceived overall sleep quality was observed in the primary analysis but was not sustained in the per-protocol analysis. Findings were consistent across ITT, per-protocol, and multiple imputation analyses for the primary outcome. While the use of comfort items in this age group most likely presents little to no risk of harm, the REST trial does not support a clinically meaningful effect on sleep-related impairment over one week.

## Data Availability

All participants were children and the underlying data sets cannot be shared publicly because there is no ethical approval to do so. The analysis code and all other underlying data is available in the supporting information files or at Open Science Framework (OSF) repository: The REST (Randomised Evaluation of Sleeping with a Toy or comfort item) trial: https://doi.org/10.17605/OSF.IO/WU7Q3

## Declarations

## Acknowledgements

The authors gratefully acknowledge the children and parents who participated in The Kid’s Trial for their time and engagement. We also thank colleagues across our University, the Toto Centre Initiative, and our broader networks for promoting the study within their communities.

Artificial intelligence (ChatGPT 5.0) was used in the following two ways to help prepare this manuscript: 1) language editing, and 2) drafting R code snippets consistent with our pre-specified SAP. All methodological and analysis decisions, and interpretations of results, were made by the authors. The authors ran all analysis code, and checked it for accuracy, with revisions made as necessary, and take full responsibility for the content of the published article.

This work contributes to one of the authors’ (SL) doctoral projects.

## Ethics approval, consent to participate and consent to publish

This study received ethical approval on 16 January 2023 from the University of Galway Research Ethics Committee (REC) (Ref: 2023.02.014). The participant information leaflets approved by the Research Ethics Committee advised participants that the overall findings of The Kid’s Trial, including the REST trial, would be submitted for publication. All participants’ legal guardians acknowledged that they had read and understood the contents of the participant information leaflets when consenting to their children’s participation in The REST Trial. Individual participants are not identifiable in any reports or publications.

## CRediT authorship statement

Conceptualisation: DD, LF, SL; Data Curation: DD, LF, NT, SL; Formal Analysis: SL; Funding Acquisition: DD; Investigation: DD, SL; Methodology: DD, LF, NT, SL; Project Administration: DD, LF; NT, SL; Resources: N/A; Software: N/A; Supervision: DD, LF, NT; Validation: DD, LF, NT, SL; Visualisation: DD, LF, NT, SL; Writing- Original Draft Preparation: SL; Writing- Reviewing & Editing: DD, LF, NT, SL.

## Data availability statement

The datasets resulting from the REST trial are not available to the public. All other supporting information is publicly available in the Open Science Framework (OSF) repository: The REST (Randomised Evaluation of Sleeping with a Toy or comfort item) trial: https://doi.org/10.17605/OSF.IO/WU7Q3 [28] under the terms of the Creative Commons Zero “No rights reserved” data waiver, CC0 1.0 Public domain dedication.

## Supporting Information

S1 CONSORT Checklist

S2 REST Trial Outcome Questionnaires:

S2a_Outcome Questionnaire_Control Group
S2b_Outcome Questionnaire_Intervention Group

S3 SAP (Statistical Analysis Plan)

S4 R Markdown Scripts

S5 Observed vs. Multiple Imputation (MI) adjusted mean difference in SQS at Post-Test

